# Central-positive complexes in ECT-induced seizures: Evidence for thalamocortical mechanisms

**DOI:** 10.1101/2020.04.28.20072520

**Authors:** Emma R. Huels, L. Brian Hickman, ShiNung Ching, Eric J. Lenze, Nuri B. Farber, Michael S. Avidan, R. Edward Hogan, Ben Julian A. Palanca

## Abstract

Electroconvulsive therapy (ECT) relies on the electrical induction of generalized seizures to treat major depressive disorder and other psychiatric illnesses. These planned procedures provide a clinically relevant model system for studying neurophysiologic characteristics of generalized seizures. We recently described novel central-positive complexes (CPCs), which were observed during ECT-induced seizures as generalized, high-amplitude waveforms with maximum positive voltage over the vertex. Here, we performed a systematic characterization of 6,928 CPC ictal waveforms recorded in 11 patients undergoing right unilateral (RUL) ECT. Analyses of high-density 65-electrode EEG recordings during these 50 seizures allowed evaluation of these CPCs across temporal, spatial, and spectral domains. Peak-amplitude CPC scalp topology was consistent across seizures, showing maximal positive polarity over the midline fronto-central region and maximal negative polarity over the suborbital regions. Total duration of CPCs positively correlated with the time required for return of responsiveness after ECT treatment (*r* = 0.39, *p* = 0.005). The rate of CPCs showed a frequency decline consistent with an exponential decay (median 0.032 (IQR 0.053) complexes/second). Gamma band (30-80 Hz) oscillations correlated with the peak amplitude of CPCs, which was also reproducible across seizures, with band power declining over time (*r* = −0.32, *p* < 10^−7^). The sources of these peak potentials were localized to the bilateral medial thalamus and cingulate cortical regions. Our findings demonstrated CPC characteristics that were invariant to participant, stimulus charge, time, and agent used to induce general anesthesia during the procedure. Consistent with ictal waveforms of other generalized epilepsy syndromes, CPCs showed topographic distribution over the fronto-central regions, predictable intra-seizure frequency decline, and correlation with gamma-range frequencies. Furthermore, source localization to the medial thalamus was consistent with underlying thalamocortical pathophysiology, as established in generalized epilepsy syndromes. The consistency and reproducibility of CPCs offers a new avenue for studying the dynamics of seizure activity and thalamocortical networks.

## Introduction

Electroconvulsive therapy (ECT) employs the deliberate induction of generalized seizures through the delivery of current to the scalp. ECT is conducted using general anesthesia and muscle paralysis, allowing for predictable seizure timing while minimizing potentially harmful tonic-clonic activity. These aspects confer the potential of ECT as a model system for leveraging electroencephalography (EEG) to examine circuits underlying generalized seizure evolution, termination, and associated cognitive outcomes (McNally and Blumenfeld, 2004).

The EEG evolves during the progression of generalized ECT-induced seizures. Weiner described five phases of ECT-induced ictal EEG: preictal; epileptic recruiting activity, polyspike activity, polyspike and slow wave activity, and termination (Weiner, 1982). Alternatively, Brumback and Staton described three phases of ECT-induced seizures (Brumback and Staton, 1982). Phase I was initial rhythmic beta (14–22 Hz) activity, followed by arrhythmic, lateralized polyspike activity in Phase II and rhythmic 2.5 to 3.5 Hz spike/polyspike-wave activity in Phase III. We recently further delineated these Phase III generalized EEG discharges, demonstrating that generalized, high-amplitude, central-positive complexes (CPCs) emerged during Phase III of ECT-induced seizures (Hogan *et al*., 2019). Despite variability in frequency and amplitude of CPCs, these complexes typically evolved from 4.0 to 1.5 Hz in frequency and decreased in amplitude throughout seizure progression. The spatiotemporal features and significance of these CPCs remain unclear.

In other models of epileptic seizures, spike wave discharges show synchronized firing of the neural elements in an extended thalamocortical network (Huguenard, 2019). Generalized seizure waveforms, such as in absence and juvenile myoclonic epilepsy, show topographic maximal amplitude distribution over the fronto-central regions, predictable intra-seizure frequency decline, and coupling with gamma-range frequencies (Benedek *et al*., 2016). Given the previously described properties of CPCs as high-amplitude EEG waveforms with well-defined positive and negative electrical potentials over the scalp and suborbital surfaces (Hogan *et al*., 2019), EEG source imaging with high-density EEG recordings offers the possibility of identifying the generating source of the electrical potentials (Kaiboriboon *et al*., 2012). While high-density EEG has allowed for characterization of the spatiotemporal dynamics of epileptic seizures (Lantz *et al*., 2003a; Holmes, 2008; Feyissa *et al*., 2017), this approach has not been employed to characterize the evolution of seizures induced by ECT.

Thus, we recorded high-density 65-electrode EEG recordings to provide a systematic understanding of these CPCs across temporal, spatial, and spectral domains. We carried out a quantitative characterization of 6,928 CPC ictal waveforms recorded from 50 seizures induced in 11 patients. Our findings provide insight into the pathophysiology of generalized seizures, as well as the recovery of consciousness. Based on their characteristics, CPCs may be important in understanding the thalamocortical circuitry underlying ECT-induced generalized seizures.

## Materials and methods

### Ethics

This is a pre-specified substudy of an interventional investigation on the recovery of cognitive function following electroconvulsive therapy (NCT02761330). All procedures were approved by the Human Research Protection Office at the Washington University School of Medicine and detailed in a previously published protocol (Palanca *et al*., 2018).

### Participants

Patients were recruited at Barnes-Jewish Hospital (St. Louis, MO) prior to initiation of right unilateral (RUL) ECT for treatment-resistant non-psychotic unipolar or bipolar depression. Participants were at least 18 years of age, fluent in English, and able to provide written informed consent. Individuals were ineligible if they had: 1) cognitive impairment sufficient to impair testing prior to ECT, 2) a diagnosis of schizophrenia or schizoaffective disorder, 3) blindness or deafness, or 4) an inadequate seizure during etomidate general anesthesia, defined as bilateral spike-and-wave complexes present for less than 10 seconds.

Individuals who discontinued study participation prior to completing the first week of seizures (*n* = 3) or provided insufficient ictal recordings (*n* = 1) were excluded from analyses. **Supplemental Table 1** includes demographic information for the remaining 11 participants who contributed a total of 50 generalized seizures for analyses.

### Study design

Patients were scheduled for a dose-charge titration session under etomidate general anesthesia (0.2 mg/kg) to determine the charge necessary to induce an adequate generalized seizure. As ECT is performed under general anesthesia through agents with diverse mechanisms and different neurophysiologic signatures, we randomized participants to the order of three anesthetic regimens. These included: ketamine (2-2.5 mg/kg bolus) with ECT, etomidate (0.2 mg/kg bolus) with ECT, and ketamine (2-2.5 mg/kg bolus) with sham-ECT. The two anesthetic regimens provided a contrast between a low-dose anesthetic with a primary mechanism of GABA agonism (etomidate) vs. a high-dose of an NMDA antagonist (ketamine). The specific order of study conditions was randomly assigned. These six treatments occurred over a two-week period, with the exception of one individual who had a week delay between two sessions, due to illness. ECT stimulation parameters were per protocol (Palanca *et al*., 2018) and the same RUL ECT charge was used during all study treatment sessions (six times the charge needed to produce a seizure during the initial dose-charge titration session). Patients and data analysts were blinded to the anesthetic type and whether ECT charge was delivered. The sham-ECT + ketamine sessions will not be discussed in this manuscript, as no seizures were induced in that condition.

### EEG acquisition

Each study session included EEG acquisition before, during, and after treatment. A 65-electrode Geodesics Sensor Net (Electrical Geodesics, Inc./Phillips Eugene, OR, USA) was modified to allow placement of the superior stimulation electrode, with a 4-6 cm window just anterolateral to the vertex. Sensor electrodes affected by cap modification (E1, E4 and E54) required signal interpolation following data acquisition (**Figure 1A-B**). Caps were sized according to patient head circumference. EEG sensor chambers were filled with Elefix electrode paste (Nihon Kohden America, Inc., Irvine, CA, USA) to maintain conductivity with the face and scalp. EEG recordings were acquired at 500 Hz, using a Net Amps 400 amplifier and a Late 2012 Mac Pro Workstation (Apple Cupertino, CA, USA) running Net Station version 5.0 (Electrical Geodesics, Inc./Phillips Eugene, OR, USA). EEG signals were synchronized with video acquired using an Axis P3364LV network camera (Axis Communications, Lund, Sweden).

**Figure 1.**
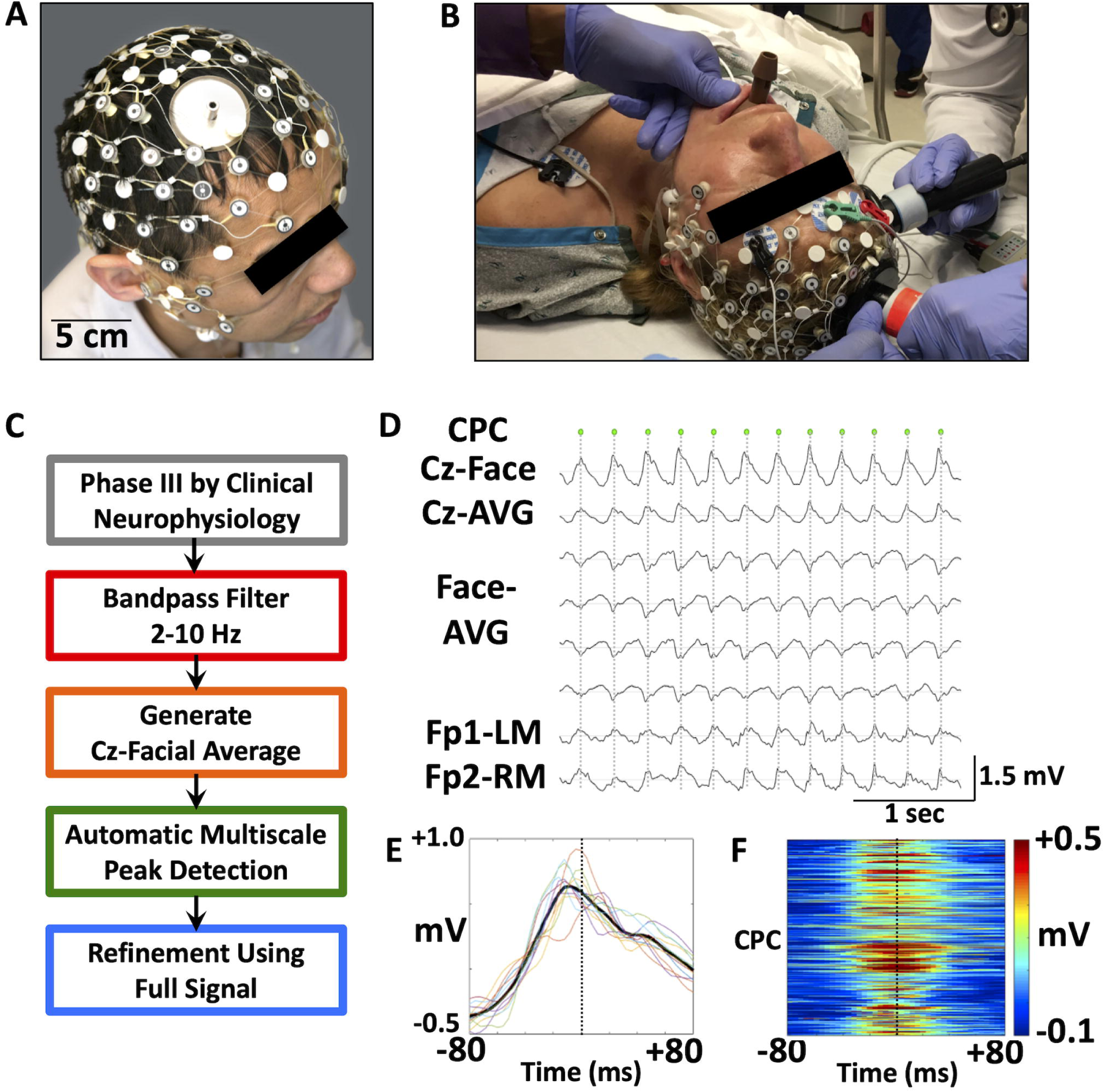
A novel system for characterizing central-positive complexes induced by right unilateral electroconvulsive therapy (RUL ECT). **(A-B)** A modified high-density 65-electrode EEG net allows signal acquisition throughout RUL ECT. EEG electrodes are positioned to permit transcutaneous charge delivery via stimulation paddles. High-density EEG signals are recorded concurrent with clinical frontomastoid EEG recordings. **(C)** A novel automated algorithm is utilized to detect central-positive complexes (CPCs) within epochs of Phase III ictal activity, defined by previous EEG criteria. Complexes are detected from a 2-10 Hz bandpass filtered signal. **(D)** Detection of large amplitude CPCs (each marked by a green dot) is based on the gradient between Cz and suborbital facial electrodes (Cz-Face). By comparison, complexes are less prominent in frontomastoid (Fp1-LM and Fp2-RM) channels approximating those typically clinically monitored for induction and termination of ECT-induced seizures. **(E)** Variance in shape and timing of the 12 CPC complexes during the initial portion of Phase III (**D**) are shown in overlay of EEG signals in Cz-Face channel. Mean signal is indicated as a heavy black trace while the dotted vertical line is for time 0. **(F)** Time-voltage maps for Cz-Face EEG for each of the 6,928 CPCs, stacked (y-axis) to show variance in time (x-axis) across 11 patients and 50 seizures. The color scale to the right represents the differential amplitude of the Cz-Face channel.

Steps were undertaken to maximize acquisition of ictal EEG while maintaining safety of the patient and protection of recording equipment. During the participation of the first six individuals, the amplifier was turned off prior to charge delivery. This resulted in approximately 5-15 seconds of EEG data loss. In order to reduce the amount of data loss for the remaining participants, the cap was disconnected from the acquisition computer immediately before charge delivery and reconnected once delivery ceased. Visual inspection (REH) (Hogan *et al*., 2019) confirmed similarity of high-density EEG and recordings utilized for seizure monitoring (Thymatron IV, Somatics, LLC, Venice, FL, USA) and no significant loss of Phase III data.

### Qualitative EEG staging

Prior to quantitative analysis, all 50 seizures included for analyses were also visually inspected and processed in Net Station Waveform Tools (Electrical Geodesics, Inc./Phillips Eugene, OR, USA). Data were preprocessed through bandpass filtering at 1-100 Hz. Channels with excessive noise were interpolated. Signals were then recomputed to the average reference. Staging was performed in Persyst software (Persyst, Solana Beach, CA, USA) by an experienced epileptologist (REH), according to previously established criteria (Brumback and Staton, 1982). Phase I was defined as 14-22 Hz rhythmic beta activity, followed by arrhythmic polyspike activity in Phase II and rhythmic 2.5/3.5 Hz sharp complex activity in Phase III (Brumback and Staton, 1982). The duration and overall description of each phase, total seizure duration, and patterns of seizure termination were noted. Additionally, seizures were evaluated for the presence and duration of central-positive complexes (CPCs) previously described through clinical readings and scalp surface topographic maps of five patients in this dataset (Hogan *et al*., 2019).

### EEG preprocessing

All ictal EEG tracings were visually inspected for excessive artefact (ERH, LBH) in Net Station Review (Electrical Geodesics, Inc./Phillips Eugene, OR, USA). EEG channels were excluded (median 6 channels/recording, IQR of 4 channels/recording) if they exhibited excessive artefact. The EEG recordings were imported into EEGLab (Delorme and Makeig, 2004) and processed using custom-written functions in MATLAB 2018a (Mathworks, Natick MA). Data were bandpass filtered at 1-100 Hz and then downsampled to 250 Hz. A 60 Hz notch filter was employed to reduce electrical artefact. Signals at previously rejected electrode locations were estimated using spherical spline interpolation. Finally, signals were re-referenced to the global average.

### Quantitative detection of CPCs

Quantitative analyses focused on characterizing CPCs over time, frequency, and space. We have qualitatively described these large-amplitude complexes that occur during Phase III (Hogan *et al*., 2019). An automated algorithm was generated for quantifying CPCs (**Figure 1C-D**). Complexes were detected based on a priori assumptions of a superior-inferior gradient via generation of the difference between Cz and the average signal among suborbital facial electrodes (Geodesics Sensor Net E61-E64). This signal was bandpass filtered between 2-10 Hz to minimize noise and drift (Scherg *et al*., 1999). Given the fluctuating amplitude of the ictal EEG and variance in peak amplitude across seizures and individuals, Automatic Multiscale-based Peak Detection (AMPD) (Scholkmann *et al*., 2012) was applied to detect complexes in the filtered 2-10 Hz Cz-suborbital facial electrode averaged signal. Following this initial localization of CPCs, the time of the peak CPC voltage was determined from the maximum voltage of the broadband (1-100 Hz) signal, ascertained within the time range halfway between the initial peak time and the times of the two flanking CPCs. Variability in shape of the 2-10 Hz bandpass filtered Cz-suborbital EEG signal and timing relative to the time of the peak CPC voltage are provided (**Figures 1E-F)**.

The rate of deceleration of CPCs (φ_(i,2)_) was fit to a nonlinear exponential decay model:

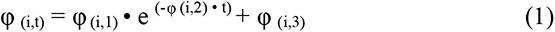

Here, the instantaneous frequency of CPCs at any time *t*, φ _(i,t)_, can be calculated from the initial frequency, φ _(i,3)_ + φ _(i,1)_, and ending frequency, φ _(i,3)_. The model was used to fit the instantaneous frequency observed by taking the inverse of the interspike interval, or time between adjacent CPCs.

### Time-frequency analysis and source estimation

Preprocessed average-referenced EEG with timing of CPCs were imported into Brainstorm (Tadel *et al*., 2011). The signals corresponding to the peak of the CPCs were subsequently used to generate topographic maps of the arithmetic mean and standard error of the mean at both an individual and group level (Tadel *et al*., 2011). Time-frequency analyses were carried out using Morlet wavelets (central frequency 1 Hz, range 1-80 Hz). The power estimates were multiplied by the frequency for 1/*f* compensation and compiled across all CPCs.

Initial steps of source estimation were carried out in a manner similar to those previously described (Stropahl *et al*., 2018). A generic head surface model with head, outer skull, and inner skull components was created based on the ICBM 152 atlas. Forward modeling relied on OpenMEEG (Kybic *et al*., 2005) boundary element method (BEM) model of scalp, skull, and brain and adaptive integration. Our source model for deep brain analysis (Attal and Schwartz, 2013) consisted of surface for bilateral cortical (15002 vertices, 2343.44 cm^2^ area, constrained), hippocampal (8404 vertices, 54.43 cm^2^ area, constrained), and thalamic (8976 vertices, 60.99 cm^2^ area, constrained) structures. Noise covariance matrices were created for each recording session based on artefact-free 1-second epochs selected from the immediate postictal period and commonly included PGES. Source estimates (surface constrained dipoles) based on dynamic statistical parametric mapping (dSPM) (Dale *et al*., 2000) were generated using the diagonal noise covariance and averaged across Phase III epochs of each individual. The approach of dSPM provides Z-score maps to gauge statistical significance. Additional exploratory source estimates were generated using standardized low resolution electromagnetic tomography (sLORETA) (Pascual-Marqui, 2002) and weighted minimum norm estimate (wMNE) (Lin *et al*., 2006). A group average was then generated across participants and projected on to the ICBM 152 structural anatomy.

### Cognitive assessments

Cognitive outcomes for the return of responsiveness were recorded. Recovery time was defined as the interval between seizure termination and the first instance at which the patient demonstrated compliance to auditory command following seizure termination. At 30-second intervals, one of two standardized auditory commands were played through a computer speaker placed near the participant’s head: “Squeeze your left hand twice” or “Squeeze your right hand twice.” As a squeeze bulb was present in each hand, compliance was defined by successful squeezing of the bulb in the correct hand. The order of these directives was randomized. The assessor of this interval was present during data acquisition but was blinded to the calculated duration of CPCs for the corresponding seizure.

### Statistical analysis

Wilcoxon signed-rank tests were used to assess differences in medians for paired samples. In instances of repeated measures (i.e. two ketamine or etomidate sessions per patient), the measurement of interest was averaged between the repeated sessions and the resulting average was used for comparison. The relationship between CPC characteristics and time were assessed using Pearson correlation coefficients.

### Data availability

All data and analysis scripts will be made available based on reasonable requests of the authors.

## Results

### High-amplitude CPCs dominate right unilateral ECT-induced seizures, irrespective of stimulus charge and anesthetic

We first evaluated ictal complexes using previously defined staging criteria (Brumback and Staton, 1982). Phase I was present in 11 of 50 seizures (median duration 5 seconds; IQR 5.5 seconds), Phase II was observed in 37 of 50 seizures (median duration 4 seconds; IQR 5.5 seconds), and Phase III was recorded in all seizures (median duration 43.5 seconds; IQR 33 seconds). Building on previous qualitative descriptions of Phase III ictal complex frequency (Brumback and Staton, 1982; Weiner, 1982), we analyzed CPCs as described in a case series (Hogan *et al*., 2019). CPCs occurred during Phase III of all recorded seizures, with a median duration of CPCs of 36.5 seconds (n = 50; IQR 24 seconds). The duration of CPCs did not significantly vary by anesthetic (*p* = 0.413, Wilcoxon signed-rank test (**Table 1**).

**Table 1.** Qualitative seizure measures. Table depicting qualitative seizure measures for all seizures induced by right unilateral electroconvulsive therapy (RUL ECT), separated by dose charge titration (with etomidate anesthesia), ketamine + RUL ECT, and etomidate + RUL ECT.

|  | Dose Titration<br>Etomidate<br>(n = 9) | ECT +<br>Etomidate<br>(n=21) | ECT +<br>Ketamine<br>(n=20) |
| --- | --- | --- | --- |
| Central Seizure Duration (s)<br>Median (IQR) | 77 (68) | 60 (38) | 49.5 (26.5) |
| Phase III Duration (s)<br>Median (IQR) | 36 (20) | 50 (36) | 41 (28.5) |
| CPC Duration (s)<br>Median (IQR) | 34 (22.8) | 45 (24.1) | 35.8 (21.5) |

To better characterize the spatiotemporal evolution of CPCs, we developed an automated algorithm to detect peaks along the superior-inferior axis (Cz referenced to the averaged suborbital facial electrodes). The frequency of these complexes declines over time (**Figure 2A-B**). The topographic distribution of peak amplitude for individual CPCs across the scalp over the course of Phase III is shown for a single participant (**Figure 2B**). Across each seizure, CPCs, as referenced to the global average, consistently showed surface polarity with peak positive amplitude over the fronto-central electrodes and peak negative amplitude inferiorly, particularly over the suborbital electrodes (**Figure 2B)**. Polarity and surface morphology remained consistent at a group level (**Figure 2C)**. The topography is invariant of stimulation charge dose or anesthetic (**Supplemental Figure 1A-C**).

**Figure 2.**
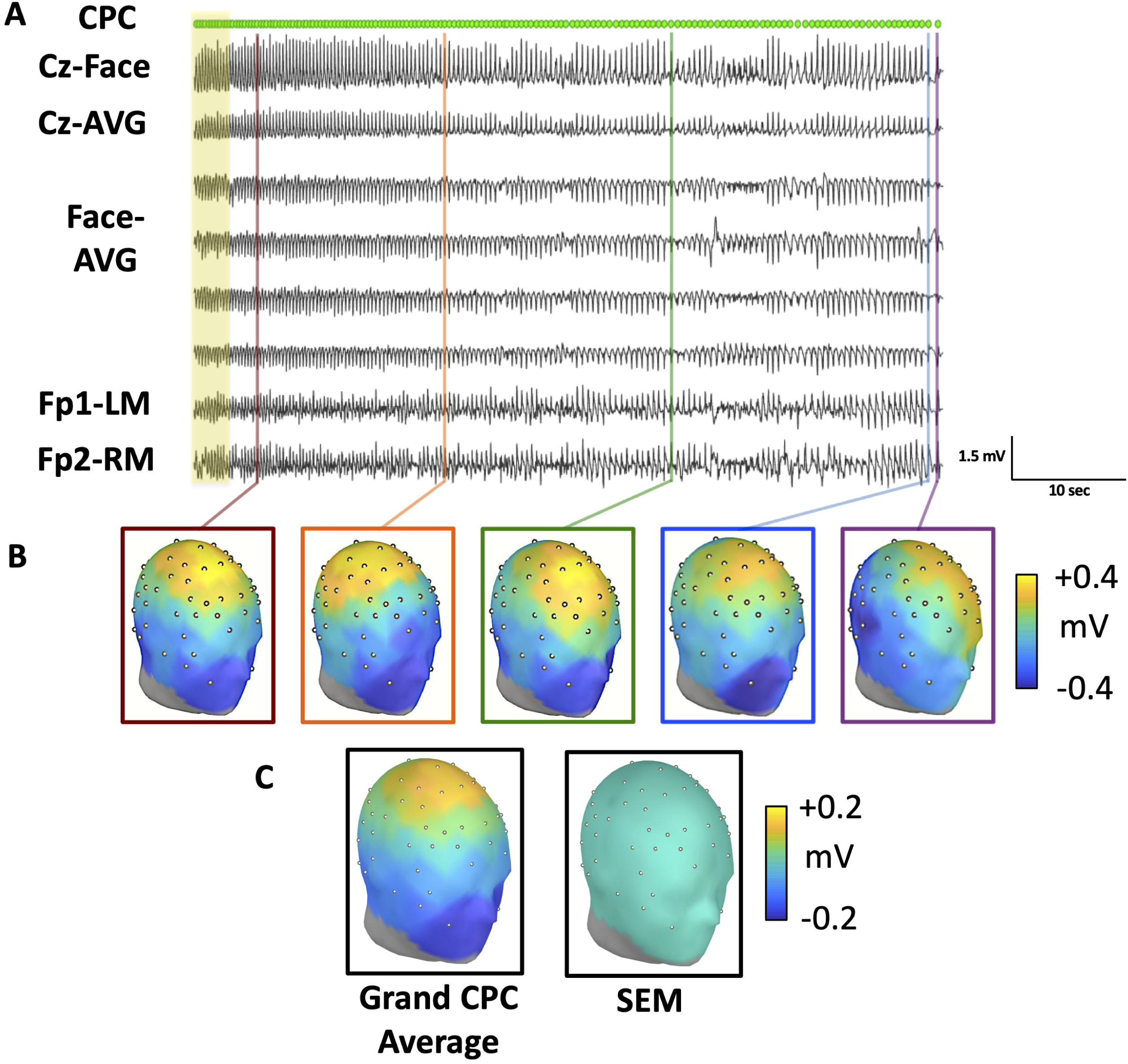
Consistency in CPC EEG scalp topography across time, ECT stimulus magnitude and anesthetic regimen. **(A)** CPCs detected during the evolution of Phase III activity during an ECT session performed under ketamine general anesthesia. Data previously shown in **1D** are highlighted in yellow. All topographic renderings represent peak amplitudes of CPCs, using a global average reference, with time sampling of 4 ms. **(B)** Topographic maps of different peak voltage of the CPC during evolution of the single seizure as depicted in **2A**. Times of sampling are indicated by corresponding colors of vertical lines in **2A** and boxes in **2B**. Scalp distribution of peak topographic amplitudes for CPCs are relatively consistent during the course of the seizure, despite variance in signal amplitude and frequency of occurrence. **(C)** The grand average of all subjects, as well as the standard error mean of the topology of all 6,928 CPCs demonstrate consistency across individuals and conditions.

### Duration of CPCs correlates with recovery of responsiveness

To address the previously unknown relationship between CPCs and clinical outcomes, we compared the total duration of CPCs to the interval from seizure termination to the return of responsiveness (**Figure 3**). Across all treatments, recovery time following the ECT-induced seizure was correlated with longer total duration of CPCs (*r* = 0.39, *p* = 0.005, *n* = 49 seizures). Thus, CPC duration is associated with the return of responsiveness in a simple auditory-motor task.

**Figure 3.**
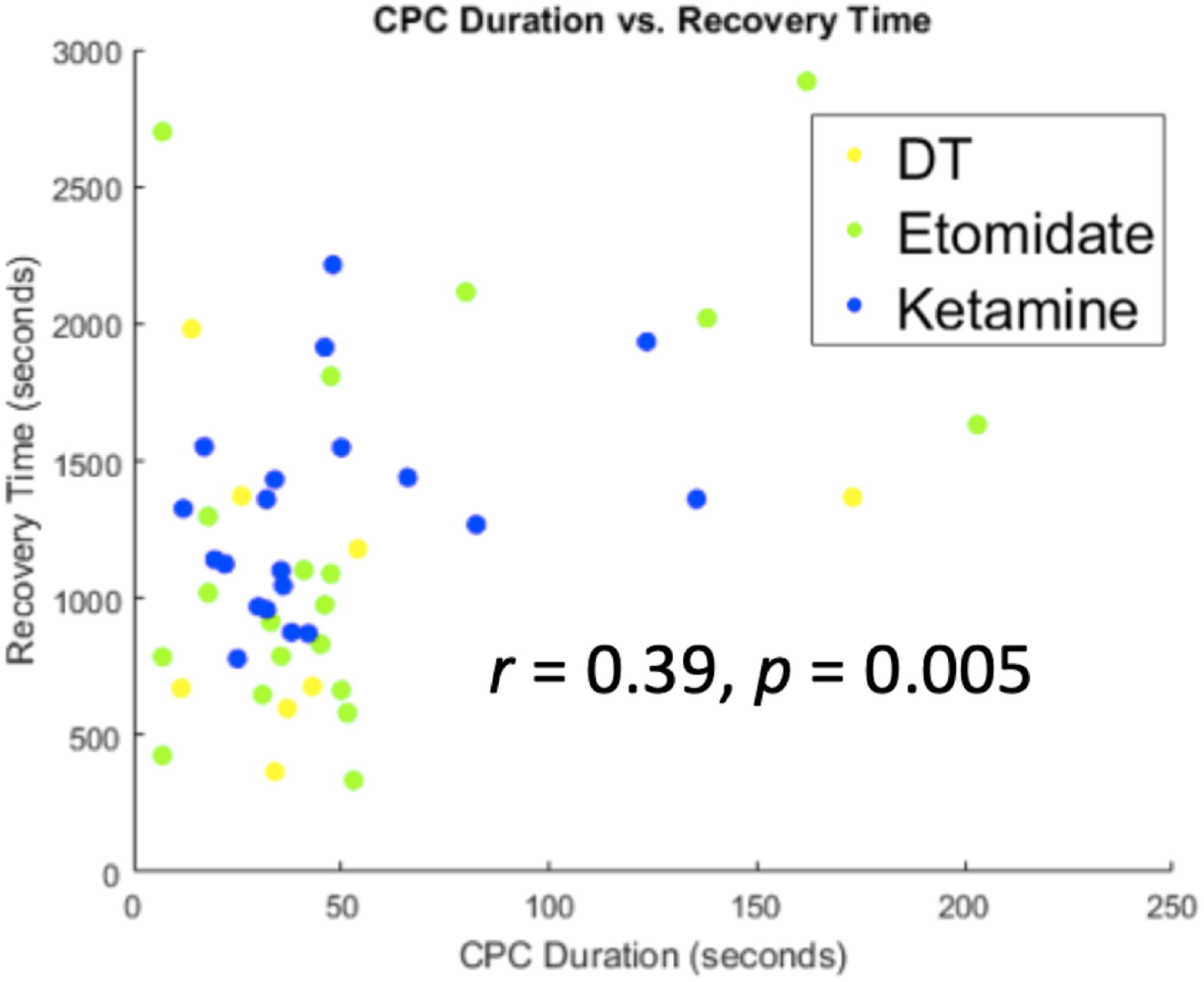
Clinical outcomes and duration of CPCs. The total duration of CPCs correlates with the interval between the end of seizure and return of responsiveness (recovery time), *r* = 0.39, *p* = 0.005. Data were pooled across dose-charge titration sessions (yellow), ECT under etomidate general anesthesia (green), and ECT under ketamine general anesthesia (blue).

### The temporal deceleration in the frequency of CPCs follows an exponential decay function

In order to assess how the rate of CPCs changes over the course of the seizure, we calculated the instantaneous frequency of CPCs over time. CPC frequency was consistently observed to be highest at the start of CPCs, with a median starting frequency of 3.93 Hz (IQR 0.73 Hz). We modeled the change in frequency over time using a damped exponential function (**Equation 1**). Examples of three modeled seizures induced under ketamine or etomidate anesthesia are shown in **Figure 4A-C**. This modeling approach fit a majority of recordings well, with a median *r^2^* value of 0.545. The median rate constant describing deceleration of CPCs for all seizures was 0.032 complexes/second, with substantial variability across seizures (IQR 0.053 complexes/second). The distribution of rate constants is illustrated in **Figure 4D**. This rate did not vary by anesthetic (ketamine: 0.049; etomidate: 0.023 complexes/second, *p* = 0.391, Wilcoxon signed-rank test). CPCs terminated at a median frequency of 2.13 Hz (IQR 0.85). The distribution of CPC frequency at termination is shown in **Figure 4E**.

**Figure 4.**
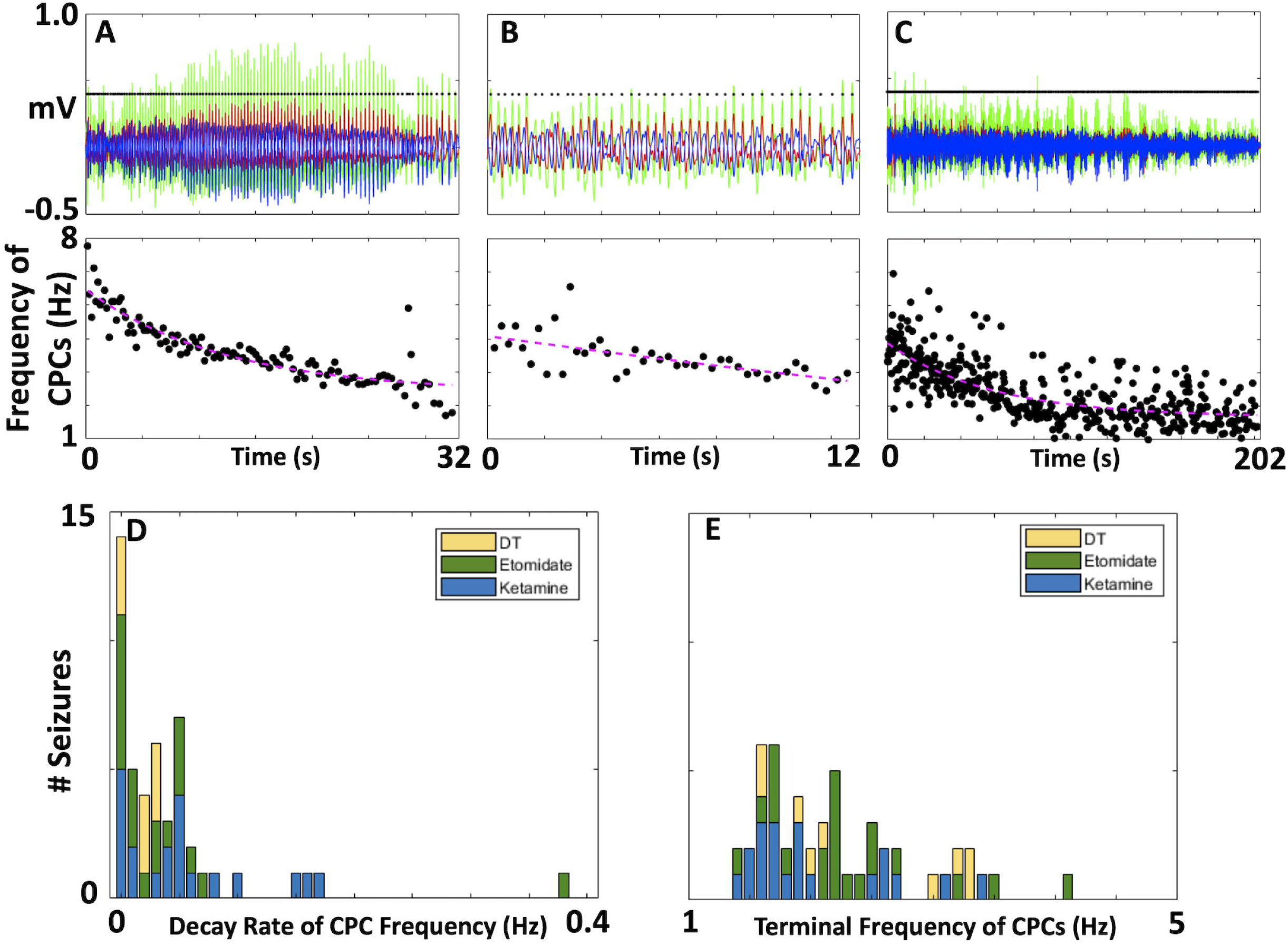
Frequency of CPC occurrence follows a damped exponential pattern without clear dependence on anesthetic regimen or stimulus magnitude. **(A-C)** EEG signals, frequencies of CPCs, and quantification of temporal decline in occurrence, using seizures from three different patients anaesthetized with ketamine (**A-B**) or etomidate (**C**). Upper panel: Overlay of EEG signals demonstrating positive potential in Cz-Avg (red), negativity in average of suborbital facial electrodes (blue), and overall high amplitude of Cz-Facial average signals (green). Times of CPCs are identified by black dots. For **(C)**, note that the density of the complexes is high such that the individual dots are not discernible. Lower panel: Instantaneous frequency of CPCs is estimated from the time interval between complexes (Black dots). These complexes are modeled to fit a damped exponential function (dashed purple) as they decay over time. **(D-E)** Modeling the evolution of CPCs during Phase III yields the rate of decline **(D)** and the frequency of CPCs at their termination **(E)**. Distributions overlap across conditions of dose-titration under etomidate (Dose-titration, yellow), ECT during etomidate (green), and ECT during ketamine (blue).

### Gamma band EEG power of CPCs declines over time

We assessed gamma band (30-80 Hz) EEG oscillations that may be associated with underlying mechanisms of seizure termination. The use of intravenous pharmacologic muscle paralytic and general anesthesia for ECT could facilitate the characterization of high frequency EEG activity during generalized seizures. High frequency (30-80 Hz) oscillatory activity was associated with CPCs (**Supplemental Figure 2**) over a wide time scale between complexes. Time-frequency analyses for an example seizure (**Figure 5A**) demonstrated both the temporal decline in CPC occurrence as well as spectral content associated with each complex. For this same seizure, total power in the gamma band (30-80 Hz) at the occurrence of each CPC negatively correlated with time (*r* = −0.53, *p* < 10^−5^, **Figure 5B**). We assessed this relationship across all seizures by calculating the correlation coefficients between band-limited power and the time of each CPC. Gamma band power was negatively correlated with time in 33 of 50 seizures (**Figure 5C**), resulting in a negative median correlation coefficient (median *r* = −0.32, IQR 0.33, *p* < 10^−7^, Wilcoxon signed-rank test). Similarly, 30 of 50 seizures had a significant decline in beta power (13-30 Hz) (**Figure 5D**). The median correlation coefficient for beta band power was less than zero (median *r* = −0.32, IQR 0.36, *p* < 10^−7^). Less consistency was noted in the delta (1-4 Hz) band (**Figure 5E**), where 9 of 50 seizures had an increase in power over time. Thirty-five seizures had a significant temporal decline in delta power, mirroring a negative correlation coefficient for delta band power (median *r* = −0.59, IQR 0.73, *p* < 10^−5^). Overall, a decline in total gamma band power over time was the most consistent finding.

**Figure 5.**
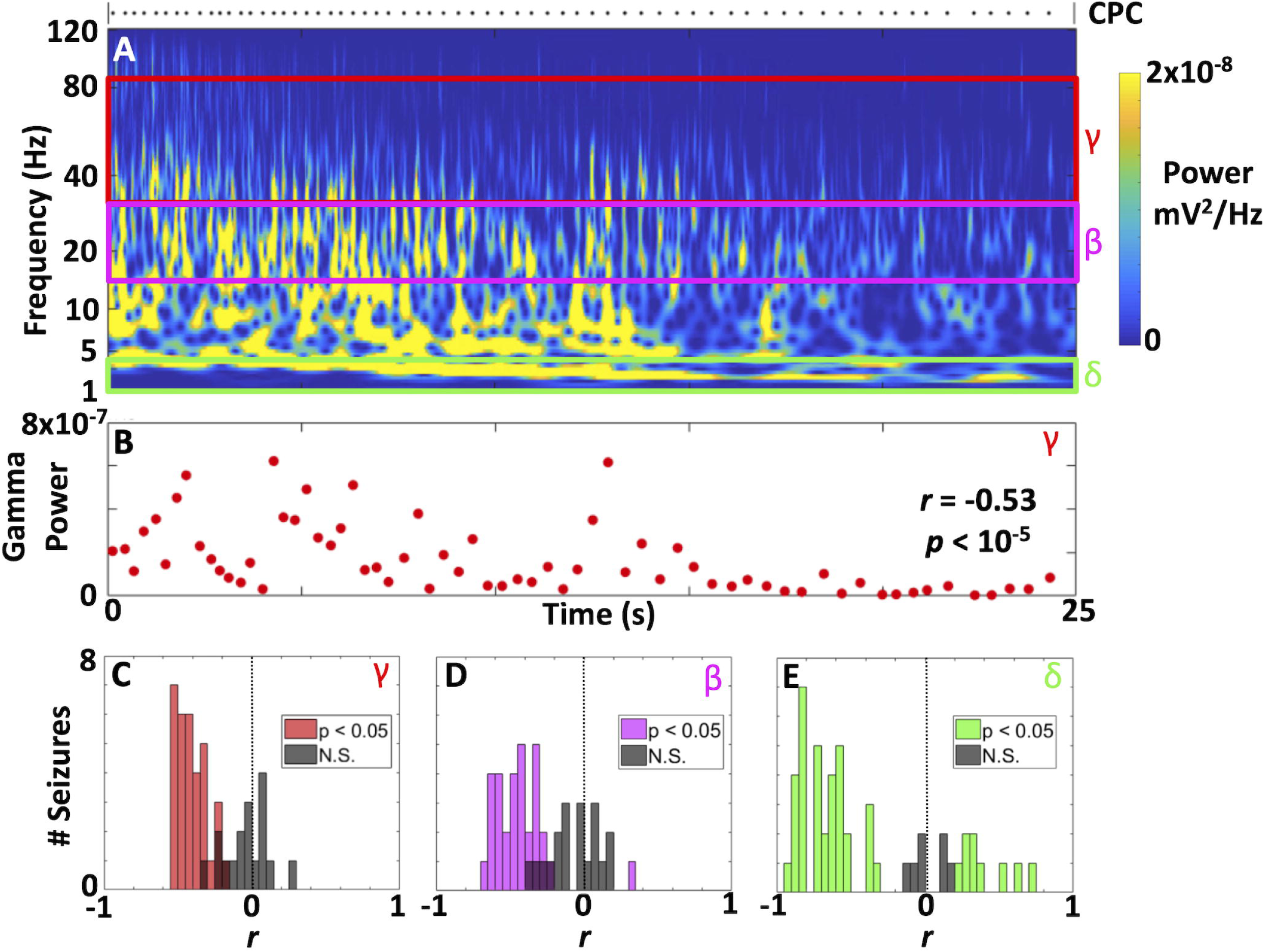
EEG spectral power of CPCs during ECT-induced seizures reflects both rate of occurrence and high frequency content. **(A-B)** Power spectrograms for Phase III ictal recordings from an exemplar patient. Deceleration in the rate of detected CPCs (black dots) is averaged in the lower frequency (2-5 Hz) ranges to show continuous signal, but shows decrease in frequency in this range. Power in harmonic multiples of CPC frequencies of occurrence are discernible within and below the alpha band (8-13 Hz). Beta (13-30 Hz) and gamma (> 30 Hz) bands also contain discernible components of EEG power attributable to CPCs. **(B)** For the same seizure in **5A**, the corresponding power in the gamma band at each CPC demonstrating a decline over time (negative slope for the correlation coefficient, *r*). (**C-E**). These correlation coefficients (as shown for a single seizure in B) can be assessed across all seizures for power in the gamma (**C**), beta (**D**), and delta (**E**) frequency bands. The coefficients are primarily negative, consistent with a decline in band-limited power over time. Non-significant coefficients are depicted in grey. Overall, the median correlation coefficients for band power were negative (gamma, *r* = −0.32; beta, *r* = −0.32; delta, *r* = −0.59; all with *p* < 10^−5^ by Wilcoxon signed-rank test).

### Central-positive complexes induced during ECT are localized to deep midline cortical and thalamic structures

Using a generic head model and three complementary approaches, we performed source estimation of the peak surface amplitude of CPCs to the bilateral surfaces of the cerebral cortex, hippocampus, and thalamus. After averaging across all individuals, the group current density map was generated. A representative Cz-facial average EEG signal, scalp topology, and source estimation for the exemplary data set of **Figure 2A-B** is provided in a **Supplementary Video**. To localize and assess statistical parametric maps of these current sources (Dale *et al*., 2000), we relied on dSPM (**Figure 6**) to provide a map of the Z-scores across the surface of these brain structures. Highest Z-scores were present across the anterior, mid, and posterior cingulate cortical regions (purple arrow), as well as the superior-midline surface of the thalamus (magenta arrow). By comparison, sLORETA (**Supplemental Figure 3A**) recapitulated group-level source estimates in the cingulate gyrus (purple arrow). Other foci were identified in the superior-midline surfaces of the thalamus (magenta arrow). In contrast to estimates from dSPM, sLORETA modeled sources to the bilateral hippocampi. Finally, weighted minimum norm estimate (wMNE, **Supplemental Figure 3B**) demonstrated foci over the anterior and mid cingulate regions (purple arrow), but also superior midline frontal cortex. Neither thalamic nor hippocampal structures were identified as sources by wMNE. Overall, source estimation showed consistent localizations across methods in localizing the peak voltage of CPCs to deep midline regions of the cingulate gyrus and superior midline thalamus.

**Figure 6.**
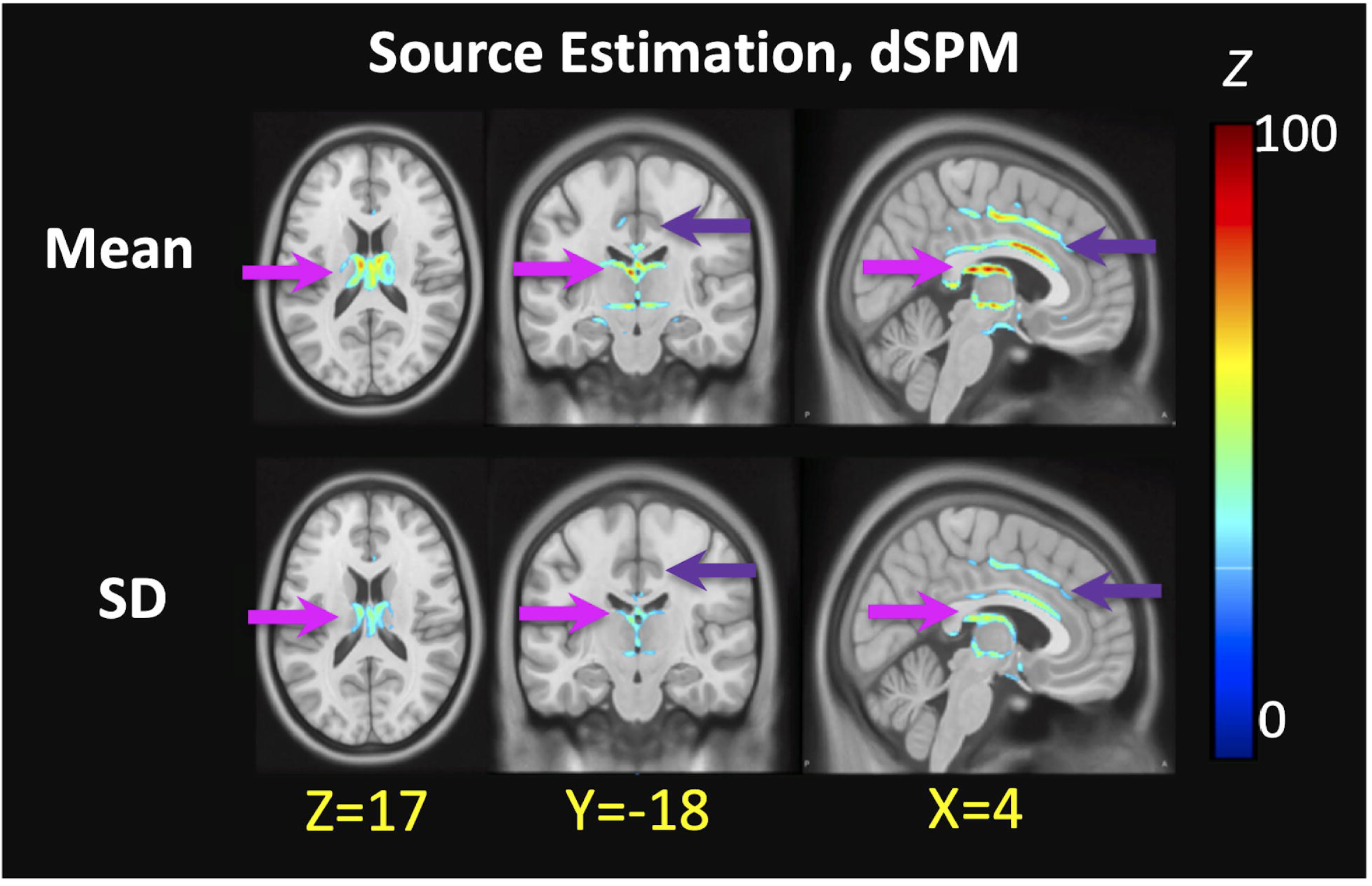
Source estimations of CPCs to deep midline cortical and subcortical structures. **(A)** Source estimation for the CPC peak potential was generated using a general head model and localization to surfaces of the cortex, hippocampus, and thalamus. Dynamic statistical parametric mapping (dSPM) allowed localized peak activity to anterior and posterior cingulate cortical regions, as well as the superior medial surface of the thalamus. Z-score estimates are shown.

## Discussion

Characterizing consistent, reproducible EEG markers for ECT-induced seizures is crucial for objective investigation of their initiation, propagation, and termination. Even within a single ECT-induced seizure, the stability of these corresponding EEG signals across space and time has been noted (Zoldi *et al*., 2000). Our current study systematically characterized the spatial, temporal, and spectral properties for 6,928 CPCs across 50 seizures. We established that these complexes are reproducible in their scalp topology across ECT stimulation charge dose and anesthetic. Additionally, we showed a predictable temporal progression of CPC occurrence during the course of these seizures. The duration of these complexes correlated with the time for return of responsiveness. These complexes were also associated with activity in the gamma frequency band. Finally, we offered preliminary analyses of surface-constrained source estimation localizing the peak signal to the superior midline thalamus and cingulate cortex. These findings have implications for the underlying pathophysiology of epileptic seizures and those induced by ECT.

### Consistency with prior characterization of EEG during ECT-induced seizures

The current report builds on prior staging criteria proposed by other investigators (Staton *et al*., 1981; Brumback and Staton, 1982; Weiner, 1982) by elucidating the morphology of robust ictal complexes typically used to document effective seizure induction in the clinical setting. Comparing these descriptions, Phase III (Brumback and Staton, 1982) matches with polyspike and slow wave activity and termination (Weiner, 1982). Weiner described the frequency of polyspike and slow-wave discharges that progress from 4-6 Hz to 1-2 Hz during the polyspike and slow wave phase, consistent with the frequency progression of CPCs (Hogan *et al*., 2019). Finally, the termination phase at the end of the seizure correlates well with our previous descriptions of seizure continuation after resolution of CPCs during some seizures. (Hogan *et al*., 2019).

In addition to the descriptions of EEG changes during ECT-induced seizures noted above, our data are also consistent with multiple studies evaluating surface topology of ECT-induced seizures. These have typically used models to document theta and delta power changes throughout the entire seizure. Sackeim and colleagues defined four patterns of delta and theta EEG topographic patterns, identifying one pattern of more prominent bifrontal delta activity as the only pattern associated with a superior clinical response following ECT (Sackeim *et al*., 1996). Luber et al. also defined 4 topographic patterns during ECT seizures, again defining one pattern with predominantly delta power over the bifrontal regions (Luber *et al*., 2000). There are significant experimental design differences between our current study defining CPCs and past studies that evaluate the entire seizure using frequency-based power spectra. However, given the delta frequency range and distribution of CPCs, their relative predominance during ECT-induced generalized seizures may explain the topographic findings in previous studies. Overall, our findings are compatible with past investigations of ictal EEG findings during ECT-induced seizures and build on their important earlier findings.

### Source localization and temporal deceleration of CPC occurrence in relationship to thalamocortical networks

A temporal decline in the rate of occurrence of CPCs and source localization of the peak potentials to the superior medial thalamic regions are consistent with the long-standing putative role of subcortical structures in the generation of generalized epileptic seizures (Norden and Blumenfeld, 2002; Huguenard, 2019). The thalamus stands out as a critically important subcortical structure in generalized epileptic seizures, serving as part of a complex, reciprocal thalamocortical network, supported by findings in both human and animal studies (Blumenfeld, 2002). Intracranial recordings in the 1950s demonstrated definitive involvement and possible initiation of generalized 3-Hz spike-and-wave seizures in the thalamus (Spiegel and Wycis, 1950; Williams, 1953). Past studies using feline and rodent epilepsy models demonstrated that the thalamus, cortex, and connecting pathways play necessary roles for the generation of typical generalized spike-and-wave seizures (Norden and Blumenfeld, 2002; Huguenard, 2019). A recent study of the Genetic Absence Epilepsy Rat of Strasbourg (GAERS) model of generalized epilepsy also showed a complex interaction between cortical and thalamic regions during generalized spike-and-wave generation. Deep somatosensory cortex demonstrated initial coupling with the ventral-postero medial thalamic nucleus (VPM) and reticular thalamic nucleus (RTN), but with subsequent coupling showing a dynamic evolution between somatosensory cortex, VPM, RTN and the posterior thalamic nucleus (Luttjohann and Pape, 2019).

In studies of human epilepsy, simultaneous EEG-functional MRI studies strongly implicate thalamic involvement in generation of generalized spike-and-wave discharges. Overall, these studies show that blood oxygenation level-dependent (BOLD) functional MRI signal is increased in the thalamus and decreased in a network of cortical regions (typically the default-mode network (Raichle *et al*., 2001)) during generalized spike-and-wave discharges, with similar findings across different generalized epilepsy syndromes (Gotman *et al*., 2005; Tangwiriyasakul *et al*., 2018). Our surface-based source modeling techniques do not allow definitive localization to individual thalamic nuclei, but the superior midline regions of the thalamus are consistent with involvement of the intralaminar and medial thalamic nuclei, both of which form extensive corticothalamocortical connections (Saalmann, 2014). These nuclei may serve an important role in generalized EEG discharges given that the centromedian nucleus of the thalamus has served as the target for electrical stimulation treatment of epileptic seizures (Fisher *et al*., 1992), with demonstrated relief from refractory epilepsy (Fisher *et al*., 2010).

Our source localization findings also support involvement of cingulate and midline frontal cortical regions during CPCs. Findings indicative of frontal cortical involvement correlate with scalp topographic findings of high amplitude activity over the superior midline frontal region and are consistent with previous data on extensive corticothalamic network interactions during generalized seizures. While there is less published data on the involvement of the cingulate cortex in the generation of generalized EEG discharges, others have demonstrated involvement of the thalami, precuneus and cingulate cortex in the etiology of generalized spike-and-wave discharges of absence epilepsy (Youssofzadeh *et al*., 2018). Detailed studies of cortical network involvement show that while thalamocortical interactions play a major role in the generation of generalized seizures, there are also complex, dynamic interactions with other brain regions in broad temporal relation to the electrographic generalized event (Bai *et al*., 2010; Tangwiriyasakul *et al*., 2018). Overall, our findings from CPC source estimation support past theories of thalamocortical involvement in generalized EEG ictal complexes for ECT-induced seizures.

In leveraging a novel automated algorithm to detect changes in CPC occurrence over time, our findings are consistent with previous reports. The method of characterizing the frequency of occurrence of CPCs over time extends previous approaches describing the temporal changes in the dominant frequency of these complexes as seizures progress (Staton *et al*., 1988). These approaches relied on linear and stepwise linear regression. Our use of damped exponentials can account for both slow (Staton *et al*., 1988) or rapid decelerations observed in the initial frequency progression of CPCs during different seizures. Overall, our approach to model the temporal dynamics of CPCs accounts for early descriptions of the temporal changes in the frequency of spike-and-wave discharges during progression of absence seizures (Williams, 1953) and complements other approaches based on frequency of clonic movements (Bauer *et al*., 2017) or ictal complexes (Jirsa *et al*., 2014) during generalized tonic-clonic epileptic seizures.

The neural underpinnings for the deceleration in the frequency of CPCs likely involve multiple mechanisms at different spatiotemporal scales and neurotransmitter systems. These mechanisms could limit both spatial propagation of seizure activity and frequency of complexes, complementing insights of large-scale coordinated networks of brain regions afforded through other seizure models (Lado and Moshe, 2008; Smith *et al*., 2016). For example, thalamocortical networks, previously discussed above, are likely responsible for negative feedback loops and intracortical inhibitory control mechanisms, particularly in midline frontal cortex (Gotman *et al*., 2005; Huguenard, 2019). The temporal decline in the frequency of CPCs may arise, in part, from thalamic activation of inhibitory cortical interneurons over excitatory pyramidal neurons (Chang and Shyu, 2014). The augmented inhibitory tone within cortical regions may then lead to suppression of cortical pyramidal neurons involved in both intracortical pathways for focal ictal wavefront spreading (Pinto *et al*., 2005; Liou *et al*., 2018) and reciprocal activation of thalamic nuclei.

### Comparison between electrographic characteristics of CPCs and absence seizure spike-and-wave discharges

There are similar EEG properties between absence 3 Hz spike-and-wave discharges and CPCs. Both 3 Hz generalized spike-and-wave discharges and CPCs are high amplitude scalp EEG complexes, with voltage maxima in the fronto-central midline and frequency evolution of high-amplitude complexes from higher to lower frequencies during the ictal event (Seneviratne *et al*., 2017). Absence epilepsy waveforms typically evolve from 3.5 to 2.5 Hz during the seizure. Weir described the generalized EEG waveform associated with absence epilepsy by three components as follows: a negative spike 1 (25–50 microvolts), a positive transient, and the main negative component spike 2 (100-200 microvolts) (Weir, 1965). This spike complex is followed by a dome-shaped surface negative wave lasting 150–200 ms. Compared to absence spike-and-wave complexes, the simpler morphology of CPCs shows a large amplitude *positive* (rather than negative) potential over the fronto-central midline. The simpler waveform of CPCs may allow a more straightforward objective analysis of its components, including modeling for source localization.

### Spectral content associated with CPCs

Spectral analyses have dominated the majority of early research involving ECT, with previous studies revealing predominantly delta activity during seizures and much smaller peaks in the gamma band (Wahlund *et al*., 2009). Gamma oscillations have become a predominant area of interest in multiple fields of study, including epilepsy (Benedek *et al*., 2016) and depression (Fitzgerald and Watson, 2018). Using cross-coherence analysis of gamma activity, Benedek et al. showed that generalized spike wave discharges in idiopathic generalized epilepsy were associated with an abrupt increase in gamma range activity, peaking at 30-60 Hz. The rise in gamma coherence decreased before cessation of the associated generalized spike wave discharges. (Benedek *et al*., 2016). We have shown associations of delta, beta, and gamma power spectra in relationship to CPCs (**Figure 5**). Throughout the course of the seizure, there remains an association of concomitant increased gamma power with CPCs. Representative EEG samples illustrate fluctuation of gamma activity (30-80 Hz) with CPCs, which by visual inspection shows a concomitant increase in gamma activity with these complexes (**Supplemental Figure 2**). This is similar to the fluctuations of 30-60 Hz activity with 3-4 Hz spike wave discharges in idiopathic generalized epilepsy (Benedek and Lauritzen, 2016). Future studies may employ coherence or phase synchrony analysis to better define the relationship of CPCs with higher frequency activity. However, our initial findings of associated gamma activity with CPCs offer further evidence of a probable thalamocortical mechanism for CPCs, given that gamma rhythms may represent activity synchronizing cortical and thalamic generators during generalized seizures (Benedek *et al*., 2016).

### Relationship of ECT-induced seizures, return of responsiveness and epileptic seizures

Impaired consciousness associated with epileptic seizures is an important clinical issue. Due to the associated sudden change in awareness of surroundings and inability to control behavior, disruptions in consciousness during epileptic seizures are a major source of morbidity and mortality for patients with epilepsy (Blumenfeld, 2012). Despite the clinical importance of consciousness in epilepsy, objective clinical study of changes in consciousness has been limited by numerous factors, including involvement of different brain regions within specific seizure types, unpredictable occurrence, and wide variability of severity of seizures (Gloor, 1986). However, absence epileptic seizures are amenable to objective study of associated changes in consciousness, given the consistent EEG manifestation of generalized spike-and-wave discharges, ability to induce seizures with hyperventilation, and recording of ictal scalp EEG with relatively little associated movement artefacts. This has resulted in many past studies correlating clinical manifestations of absence epileptic seizures with cognitive testing, functional neuroimaging changes, and EEG findings (Blumenfeld, 2012).

Similarly, patients undergoing ECT provide another population with homogenous seizure type, minimal associated motion during seizures, and predictable seizure timing. We demonstrated a significant correlation between the duration of CPCs and the time for return of responsiveness after ECT. This finding supports the importance of the electrographic properties of seizures over other factors during ECT, such as associated use of anesthetic agents, in the return of neurological function after ECT. Correlating more specific postictal EEG and cognitive changes with electrographic features of ECT-induced seizures may offer further insights into changes in consciousness related to seizures.

Another important consideration in comparing ECT-induced seizures with epileptic seizures is the possible effect of stimulation montage for seizure induction (RUL, bifrontal, or bitemporal). Past studies comparing ECT-induced and epileptic seizures equate bilateral frontotemporal ECT stimulation with typical generalized tonic–clonic seizures, resembling generalized seizures in patients with epilepsy in both clinical and EEG manifestations (Association, 2000). Seizures induced by right unilateral ECT resemble spontaneous focal seizures with secondarily generalized tonic clonic seizures (Kriss *et al*., 1978a; Kriss *et al*., 1978b). However, as previously discussed, most past studies employed limited EEG montages. In studies using full scalp EEG montages, both bilateral and unilateral ECT show similar generalized sharp wave changes as seizures progress (Brumback and Staton, 1982; Weiner, 1982). Further studies of high-density EEG recordings in patients undergoing bilateral ECT will be necessary to establish whether CPCs occur irrespective of stimulation montage.

### Limitations and Future Directions

EEG source modeling is a reliable technique for localization of focal epileptic seizures (Megevand and Seeck, 2018), but there are multiple variables contributing to accurate source localization. Our source localization findings rely on an accurate source inversion, but evaluating the accuracy of source estimates is not straightforward, given there is no objective ground truth for comparison (Haufe and Ewald, 2019). Predictably, the techniques, dSPM, sLORETA, and wMNE, show different results. For example, past investigators have reported that the depth weighting biases dSPM localization toward deep sources (Hedrich *et al*., 2017), while localization to deeper cortical structures, such as the insula, are underestimated by wMNE (Hauk *et al*., 2011). This may explain the localization of peak CPC potentials to the thalamus by dSPM and sLORETA, but not wMNE. Thus, we viewed overlap among source estimates to infer localization, emphasizing dSPM modeling as it allows statistical evaluation. The high amplitude and large number of complexes (*n* = 6,928) bolsters our confidence in the accuracy of these findings.

Aside from the processing algorithm, we acknowledge that misleading localization can arise from dipole source modeling to a spatially extended area (Kobayashi *et al*., 2005). Another important issue involves the significant propagation of cerebral activity that can occur during an epileptiform discharge (Lantz *et al*., 2003b). Time point selection within the pertinent waveform is therefore important in our source modeling of CPCs. In past modeling of epileptiform discharges, investigators applied source localization to the onset, peak, or any point in between (Megevand and Seeck, 2018). In our analysis, we chose the peak of CPCs for modeling. In future studies, evaluating changes in source localization at different time points in the CPC waveform may leverage the richness of these complexes in the temporal domain. With a denser EEG array, digitized localization of electrodes, and individual-specific anatomy, future studies may pave the way for further insight into the propagation of these seizures through discrete corticolimbic or thalamocortical pathways (Holmes, 2008).

Our current analysis focuses on characterization of CPCs. However, other important potential implications for CPCs and ECT-induced ictal electrographic changes exist, including ongoing delta activity after seizures (Fink, 2004), relationship to PGES (Hogan *et al*., 2019), and association with peri-ictal cognitive changes. Additionally, CPCs may play an important role in predicting ECT treatment outcomes and provide insight into the underlying mechanisms for efficacy of ECT in treatment of neuropsychiatric disorders.

## Summary

Our quantitative analysis of 6,928 CPCs demonstrates a consistent occurrence and relatively uniform morphology in all ECT-induced generalized seizures. The large sample size, relative uniformity, and high amplitude of CPCs allows for robust statistical analysis of their characteristics. Consistent with ictal waveforms of other generalized epilepsy syndromes, CPCs showed maximal topographic distribution amplitude over the fronto-central regions, source localization to the midline thalamus and cingulate cortex, predictable intra-seizure frequency decline following an exponential decay, and concomitant amplitude fluctuations with gamma-range frequencies. Taken together, our findings support the premise that thalamocortical mechanisms underlie the generation of CPCs. The association of delay in return of responsiveness in subjects with longer duration of CPCs is clinically important, highlighting potential clinical implications of CPCs with changes in consciousness. Overall, the consistency and reproducibility of CPCs offers a valuable marker for elucidating the electrographic dynamics of generalized seizure activity and thalamocortical networks.

## Data Availability

All data and analysis scripts will be made available based on reasonable requests of the authors.

## Abbreviations

AMPD: Amplitude multiscale peak detection
BEM: Boundary element model
CPC: Central-positive complex
dSPM: Dynamical statistical parametric mapping
ECT: Electroconvulsive therapy
EEG: Electroencephalography
GABA: Gamma-aminobutyric acid
IQR: 25-75% interquartile range
wMNE: Weighted minimum norm estimate
NMDA: N-methyl-D-aspartate
RUL: Right unilateral
sLORETA: Standardized low-resolution brain electromagnetic tomography

## Acknowledgements

We thank George Mashour, M.D., Ph.D. and Max Kelz, M.D., Ph.D. for their support of the Reconstitution of Consciousness and Cognition Phase 2 (RCC2) Research Collaborative. Their suggestions strengthened this manuscript. We thank members of our team for early discussions and data collection: Dr. Mathias Basner, Hannah Maybrier, Angela Mickle, Sherry McKinnon, Jordan Oberhaus, J. Wylie Spencer, Dr. Troy Wildes, Jamila Burton, Amil Patel, Ginika Apakama, Ravi Upadhyayula, Alaira Lourido, Dr. Changwei Wei, and Chloe Stallion. Kristopher Bakos and Kathryn Vehe Pharm D provided valuable support from Barnes-Jewish Hospital Interventional Pharmacy. Indispensable assistance was provided by Drs. Michael Jarvis and Donald Bohnenkamp, as well as the nursing staff at the Barnes-Jewish Hospital ECT Suite.

## Funding

Research reported in this publication was supported by James S. McDonnell Foundation.

## Competing interests

The authors report no competing interests.

---

**Supplemental Table 1. Baseline patient demographics**.

**Supplemental Table 2. CPC incidence across seizures**. Table depicting number of CPCs for each seizure across all participants, organized by seizures induced during dose charge titration (DT) sessions, etomidate (E), or ketamine (K) during week 1 or week 2. The column labelled “Total CPCs” indicates the total number of complexes detected from each individual throughout their experimental treatment course. Grey cells indicate sessions where recordings did not occur.

**Supplemental Figure 1. Invariance of average peak CPC scalp topologies across stimulation dose charge and anesthetic regimen. (A)** Initial dose-charge titration session with etomidate general anesthesia. **(B)** ECT with etomidate general anesthesia. **(C)**. ECT with ketamine general anesthesia.

**Supplemental Figure 2. Gamma band oscillations associated with CPCs. (A)** CPCs were detected (green dots) during the course of Phase III, using Cz-Facial average signals. Initial and final 5 second intervals (yellow regions) are shown in greater detail in **(B-C)** with times of CPC peak amplitude (green vertical lines). **(D-E)** CPCs are apparent using 2-10 Hz bandpass filtering of the EEG. **(F-G)** With 30-80 Hz bandpass filtering, gamma-band oscillations associated with CPCs are evident.

**Supplemental Figure 3. Consistency of CPC source estimation**. Generic head models were utilized to localize neural sources of CPC peak positive EEG activity to surfaces of the cortex, hippocampus, and thalamus. These source estimations were averaged across all sessions and individuals. (**A)** Through standardized low-resolution electromagnetic tomography (sLORETA), CPC source estimation converged most strongly to cingulate cortical regions (purple arrow) and superficial thalamic (magenta arrows) regions. (**B)** Using weighted minimum norm estimation (wMNE), current density mapping identified cingulate cortical regions (purple arrow) in addition to superficial frontal and parietal regions.

**Supplemental Video. Consistency of CPC topology and source localization during one seizure**. Video illustrating the topology and source localization of each CPC for an entire seizure. A single channel EEG trace representing CPCs for the entire seizure in the bottom panel, with green circles above the EEG trace marking each CPC. The traveling red line marks sequential CPCs, with simultaneous projection of topographic surface maps (upper left panel) and dSPM source estimation projected to anatomic surfaces (upper right panel). CPCs consistently demonstrate maximal positive amplitude over the vertex and negative amplitude over suborbital region, while dSPM shows localization to the midline thalamus and cingulate regions.

## Notes

### Competing Interest Statement

The authors have declared no competing interest.

### Clinical Trial

NCT02761330

### Clinical Protocols

https://www.ncbi.nlm.nih.gov/pmc/articles/PMC5960711/

